# Automated Extraction of Cancer Registry Data from Pathology Reports: Comparing LLM-Based and Ontology-Driven NLP Platforms

**DOI:** 10.64898/2026.03.20.26348915

**Authors:** Thomas McPhaul, Kory Kreimeyer, Alexander Baras, Taxiarchis Botsis

## Abstract

Cancer data standardization requires converting unstructured pathology reports into structured registry variables, a mostly manual and resource-intensive task. We evaluated two automated extraction platforms: Brim Analytics, an LLM-based system that guides and orchestrates abstraction, and DeepPhe, an ontology-driven system. Using 330 pancreatic adenocarcinoma and 34 breast cancer pathology reports from Johns Hopkins Hospital, we assessed both under deployment-realistic conditions. Brim Analytics achieved high accuracy across seven registry variables in pancreatic cancer (mean 96.7%), including T stage (96.4%) and histologic grade (97.0%), with a 3.0 p.p. decline on breast cancer (mean 93.7%). DeepPhe performed comparably for N stage (96.4% pancreatic, 94.1% breast) but had notable T stage deficits (83.6% pancreatic, 70.6% breast). Per-report processing times averaged 0.9 s (Brim, pancreatic), 4.6 s (Brim, breast), 1.1 s (DeepPhe, pancreatic), and 3.5 s (DeepPhe, breast). These results indicate that LLM-based extraction can achieve high accuracy across cancer types and support automated data workflows.

## Introduction

Cancer registries constitute the foundation of population-level oncology surveillance, epidemiological research, and quality monitoring programs^1^. In the United States, over 1.9 million new cancer cases require annual registration, with pathology reports serving as the authoritative source for diagnostic, staging, and histopathologic information^2^. Current workflows are supported by automated tools but still depend heavily on manual abstraction by certified tumor registrars—a labor-intensive process requiring specialized training and introducing variability in interpretation^3^. Beyond registry work, decision-support and other oncology use cases also require large, standardized datasets derived from pathology reports and other unstructured electronic health records (EHRs) via efficient automated pipelines.

Recent advances in natural language processing (NLP), including large language models (LLMs) in the biomedical domain, offer potential solutions to these scalability challenges^4,5,6^. LLMs can interpret diverse narrative phrasing with strong contextual understanding^7^, while ontology-driven systems generate structured outputs aligned with standardized clinical concepts^8^. However, most published evaluations are conducted under idealized conditions that do not reflect deployment realities: pathology reports vary widely in structure, terminology, and formatting across institutions and time, and it remains unclear whether existing platforms can accurately extract registry-required information from unmodified clinical documents^9,10^.

Despite these technological advances, a critical gap remains between reported system performance and real-world utility in oncology and other domains. Rigorous validation requires evaluation across heterogeneous report formats, extended temporal ranges capturing evolving clinical standards, and multiple disease types to assess generalizability — essential conditions rarely investigated thoroughly or, most importantly, addressed simultaneously in existing literature. Without such evidence, the feasibility of deploying automated extraction systems at scale for cancer registry operations and clinical research remains uncertain.

To explore the ability of existing technologies to support the above tasks, we evaluated a recently developed cloud-based LLM platform and an established ontology-driven system on two solid tumor cohorts comprising pancreatic adenocarcinoma and breast cancer pathology reports sourced directly from Johns Hopkins Hospital clinical records. Our objectives were to: (a) measure extraction accuracy across key clinical variables required for cancer registry reporting and other use cases, (b) analyze patterns of extraction errors to identify systematic failure modes, (c) examine cross-disease generalizability, and (d) quantify processing times relevant to operational deployment.

## Methods

### Study Design

This retrospective study was conducted under Johns Hopkins Medicine Institutional Review Board oversight (IRB00482075). We evaluated two fundamentally different automated extraction approaches for extracting pathologic staging and tumor characteristics from unstructured pathology reports: Brim Analytics^11^, a specification-driven, variable-based LLM framework and orchestration engine, and DeepPhe^12^, an established ontology-driven information extraction system. Brim Analytics (Version 2024.10.30) is a commercial cloud-hosted platform that leverages LLMs to perform contextual interpretation of diverse clinical narrative structures. DeepPhe (Version 0.7.0) is an open-source cancer phenotype extraction system built on the Apache clinical Text Analysis and Knowledge Extraction System (cTAKES) NLP framework, employing a double-pipeline architecture that combines a domain-specific ontology with rule-based mention detection and phenotype summarization to generate structured outputs mapped to the National Cancer Institute (NCI) Thesaurus^13^. In the Brim Analytics exploration, variable development and optimization were performed exclusively on pancreatic cancer data using strict train-test separation to prevent overfitting. Finalized extraction configurations were then applied without modification to held-out pancreatic cancer validation data and an independent breast cancer cohort to evaluate cross-disease generalizability. These two datasets were also used in the DeepPhe evaluation.

We selected pancreatic cancer as the primary evaluation domain because it poses challenging abstraction requirements: complex anatomic relationships requiring precise topographic coding, frequent multifocal disease complicating margin assessment, variable histologic patterns, and evolving staging criteria across American Joint Committee on Cancer (AJCC) editions. To assess cross-disease generalizability—a key requirement for practical deployment—we also evaluated an independent breast cancer cohort without disease-specific system modifications.

### Pancreatic Cancer Dataset

Surgical pathology reports (N = 480) were obtained from patients with pancreatic adenocarcinoma who underwent surgical resection at Johns Hopkins Hospital between 2006 and 2025 and were included in a curated database maintained by the pancreatic cancer group. This 19-year timeframe captures substantial evolution in documentation practices, including progressive adoption of College of American Pathologists (CAP) synoptic templates and implementation of AJCC 8th edition staging criteria. The cohort was divided into an optimization subset (n = 150) used exclusively for Brim Analytics variable development and a validation subset (n = 330) held out until all variables were finalized.

### Breast Cancer Dataset

An independent cohort (n = 34) of breast cancer resection specimens spanning 2006–2025 was assembled to assess cross-disease variable transferability without breast-specific modifications. The patient cases in this cohort were previously reviewed by the Johns Hopkins Molecular Tumor Board that maintains a database with high-quality standardized data and provided the abstractions used in this study.

### Pathology Report Characteristics

Report formats were deliberately heterogeneous to reflect real-world documentation practices. Of the 330 pancreatic validation reports, 238 (72.1%) were predominantly narrative with free-text descriptions, while 92 (27.9%) employed structured synoptic templates in addition to some free-text content. Of the 34 breast cancer validation reports, 21 (61.8%) comprised predominantly narrative free-text descriptions, while the remaining 13 (38.2%) employed structured synoptic templates. Reports from 2009–2018 predominantly used narrative formats, while reports from 2019–2025 increasingly adopted CAP synoptic structures. All reports underwent minimal preprocessing (assembling text sections and structured synoptic data into a single data file).

### Gold Standard Development

Seven oncological variables (Table 1) were selected based on complementary requirements from multiple sources. Five variables—T stage, N stage, M stage, histologic grade, and tumor site— align with the Precision Oncology Core Data Model (Precision-DM) that supports data standardization and management at the Sidney Kimmel Comprehensive Cancer Center (SKCCC) ^14^. Margin status was included based on priorities examined in internal efforts and external collaborations, while OncoTree classification is pursued specifically for the AACR GENIE project. Gold-standard annotations for these variables were developed by a trained clinical research specialist with expertise in oncology data abstraction, following Commission on Cancer (CoC) and AJCC 8th edition guidelines. To assess annotation reliability, a stratified 10% subset (n = 33) underwent independent validation by a second trained abstractor blinded to initial annotations and automated platform outputs. Inter-rater agreement was assessed using Cohen’s κ statistic, with perfect concordance achieved across all variables (κ = 1.0).

**Table 1.**
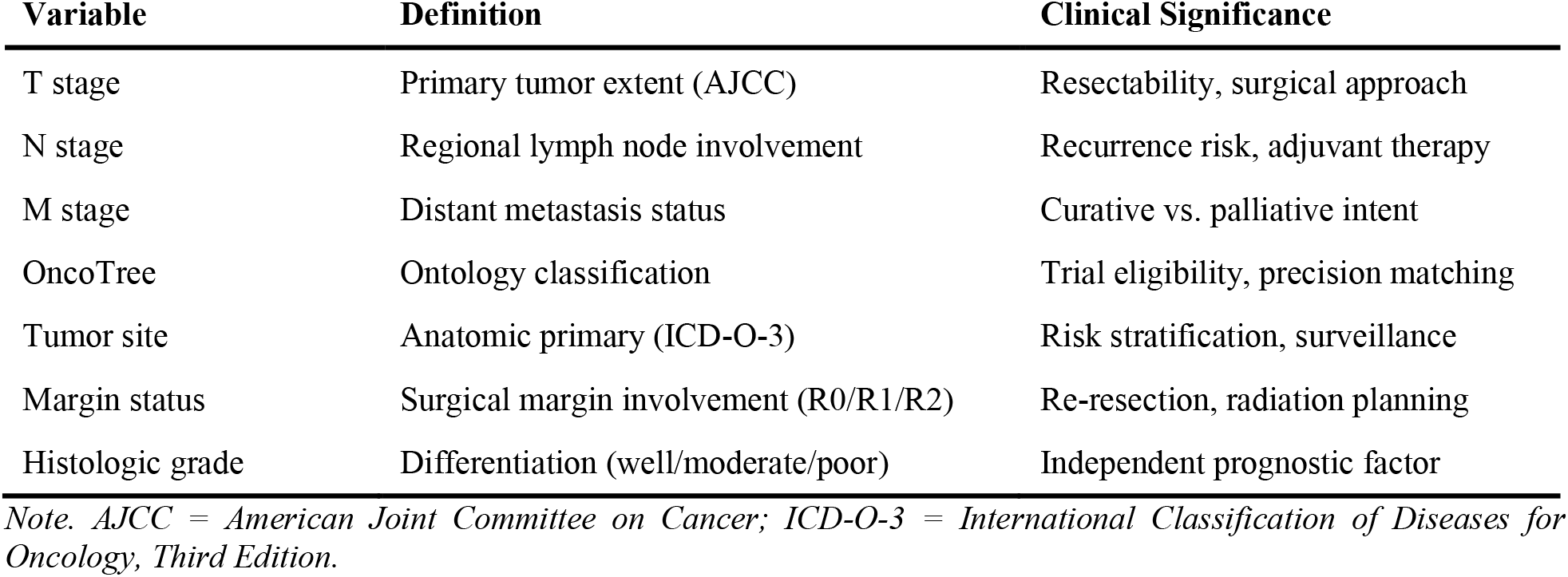
Extraction Targets and Clinical Significance.

### Clinical NLP Extraction and Validation Workflow

Pathology reports included in the SKCCC Registry were extracted from the institutional Databricks data platform that hosts this Registry. Preprocessing was limited to document assembly, whereby all constituent sections of each pathology report were concatenated into a single raw text file and assigned a study identifier to preserve document linkage, without modification, normalization, or variable-level curation of the underlying clinical content. For Brim Analytics, extraction was governed by variable definitions developed and locked on a separate development cohort prior to evaluation, grounded in established clinical standards including AJCC 8th edition staging criteria, College of American Pathologists synoptic reporting protocols, ICD-O-3, and OncoTree ontology classifications. For DeepPhe, default ontology pipeline settings were applied without any system-level customization. This design enabled direct platform comparison under equivalent input conditions, with the distinction that Brim Analytics operated under pre-specified LLM extraction instructions and DeepPhe under out-of-the-box defaults. Both systems generated case-level structured outputs containing study identifiers and extracted variable values, which were evaluated against gold-standard abstractions using confusion matrix analysis, with accuracy, precision, recall, specificity, F1 score, and Cohen’s κ calculated with 95% confidence intervals for each variable (Figure 1).

**Figure 1.**
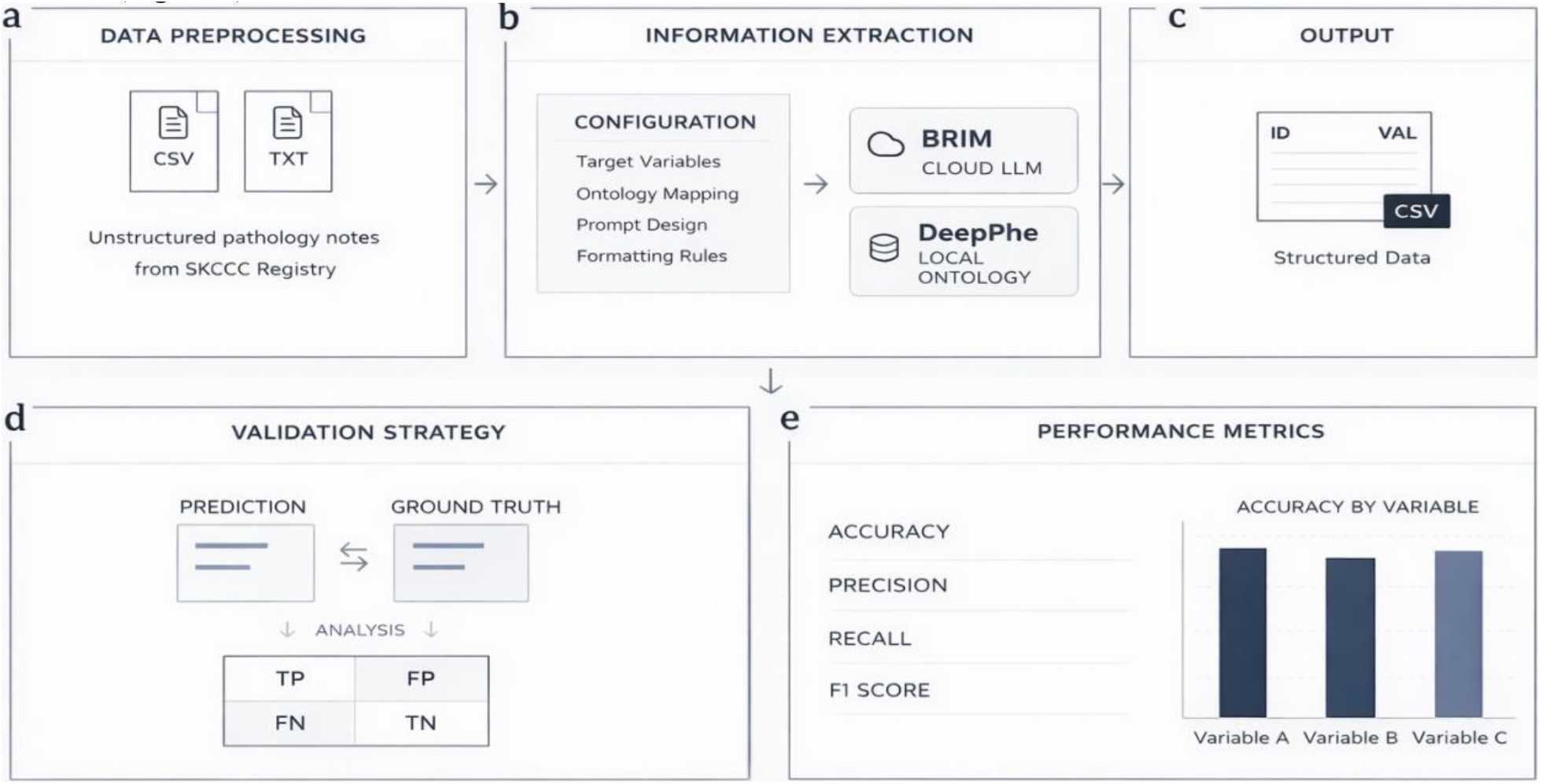
Clinical NLP extraction and validation workflow. (a) Document assembly: SKCCC reports concatenated with study IDs. (b) Extraction: Brim Analytics (guideline-based variables) and DeepPhe (default ontology). (c) Structured outputs with extracted values. (d) Validation against gold standard with train-test separation. (e) Performance metrics with 95% CIs.

### Target Variable Specification

#### Brim Analytics

Brim Analytics implements an LLM guided chart abstraction framework in which clinical domain experts explicitly define abstraction protocols—specifying what to extract, how to resolve ambiguous terminology, and when to return null values—that LLMs then execute against unstructured clinical text (Figure 2). The Brim software manages context windows, result introspection, and hallucination guardrails, among other controls. This architecture decouples rule authorship from model training, preserving transparency and auditability absent in end-to-end learned approaches. All variables were grounded in established clinical standards: Commission on Cancer registry guidelines, AJCC 8th edition staging criteria, College of American Pathologists synoptic reporting protocols, ICD-O-3, and OncoTree ontology classifications (OncoTree^15^). Critically, because extraction logic resides in explicit variable definitions rather than implicitly encoded model weights, individual extraction decisions remain fully traceable and modifiable without retraining.

**Figure 2.**
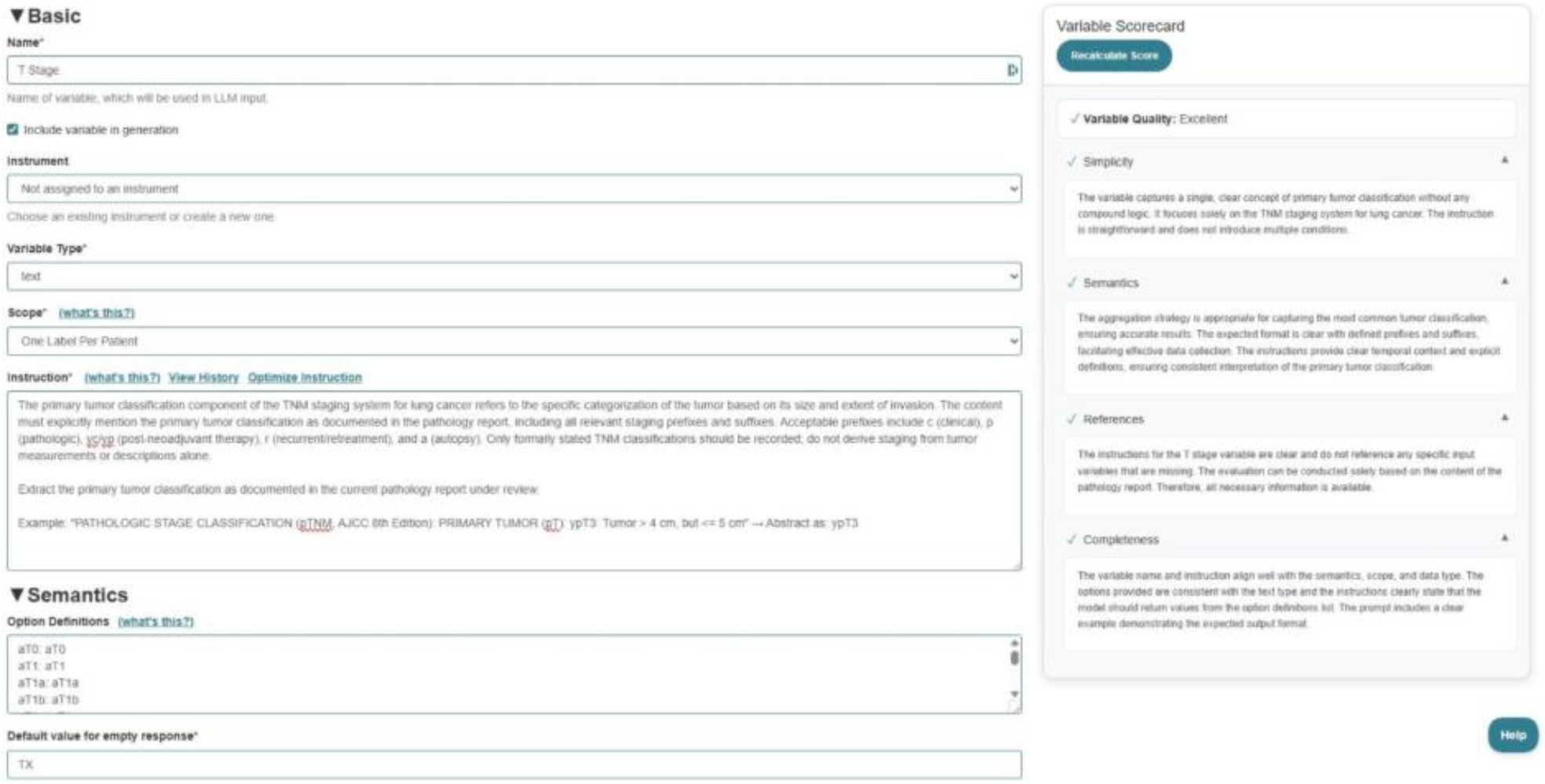
Brim Analytics Variable Configuration Interface for T Stage.

Each Brim variable comprises six structured components governing extraction behavior (Figure 2): name, variable type (e.g., boolean, integer, text), scope (i.e., the granularity of abstraction including one value per patient or one value per note), instructions, semantics, and default value. Variables were developed through a two-iteration optimization protocol applied exclusively to a pancreatic adenocarcinoma development cohort (n = 150), held strictly separate from validation datasets to ensure unbiased generalization assessment. In iteration 0, initial specifications were derived solely from published clinical guidelines without exposure to actual pathology reports, establishing baseline performance achievable through expert knowledge encoding alone. An error analysis of this initial iteration revealed four failure classes: terminology variation (for example, ‘head’ versus ‘cephalic portion’ for pancreatic subsite), ambiguous narrative phrasing, temporal inconsistencies from discordant staging across treatment timepoints, and formatting heterogeneity. In iteration 1, variable instructions were refined through clarification of ambiguous criteria, expansion of worked examples for edge cases, and comprehensive synonym mapping; all variables were then considered finalized. Finalized variables were evaluated on two independent validation cohorts: pancreatic adenocarcinoma (n = 330) to test robustness to evolving documentation practices, and breast cancer (n = 34), enabling zero-shot cross-disease transfer assessment (Figure 1).

#### DeepPhe

DeepPhe is an open-source cancer phenotype extraction system built on Apache cTAKES^13^ that employs a novel double-pipeline architecture combining a domain-specific ontology (the DeepPhe conceptual model) with rule-based mention detection and phenotype summarization^12^. The system operates on clinical documents to generate hierarchical summaries of cancer- and tumor-level characteristics across four processing levels. At the mention level, the cTAKES pipeline identifies single words or multi-word phrases corresponding to diseases, biomarkers, TNM staging elements, tumor size, procedures, and medications, along with descriptive attributes such as body location and laterality. At the document level, mentions are aggregated into structured templates using Fast Healthcare Interoperability Resources (FHIR) models to represent tumor and cancer characteristics extracted from individual reports. At the episode level, documents are classified into temporal categories (pre-diagnostic, diagnostic, treatment, or follow-up) with associated date ranges. At the phenotype level, information is integrated across all documents to generate comprehensive patient-level cancer summaries that link cancers to their primary and metastatic tumors with associated diagnostic, staging, and treatment attributes. All extracted concepts are mapped to the National Cancer Institute Thesaurus, enabling standardized representation aligned with established cancer terminology. For this study, we used DeepPhe-XN (Cancer Deep Phenotype Extraction, Translational Medicine) v0.7.0 with default pipeline configuration to extract TNM staging elements from both pancreatic and breast cancer cohorts without system-level customization.

### Statistical Analysis

Model performance was evaluated using standard classification metrics. Accuracy was defined as the proportion of correctly classified instances. Precision measured the proportion of positive predictions that were correct, while recall quantified the proportion of true positives correctly identified. The F1 score was computed as the harmonic mean of precision and recall. Cohen’s κ assessed agreement while adjusting for chance^16^. Ninety-five percent confidence intervals were calculated using Wilson score intervals^17^. Sample size and power considerations were applied to the pancreatic validation cohort (n = 330). This cohort size was selected to provide adequate precision for accuracy estimates and to detect clinically meaningful differences in between-platform performance under a paired design at a two-sided α = 0.05. The breast cancer cohort (n = 34) was included as an independent, exploratory assessment of cross-disease generalizability and was not powered for formal hypothesis testing.

## Results

Brim Analytics (v2024.10.30; GPT-4.1 mini; 150 tokens per minute rate limit) was executed on an Azure virtual machine running Red Hat Enterprise Linux 10.1 (Coughlan) with 4 logical CPU cores and 16 GB RAM. The platform processed the pancreatic cohort in 4 min 55 s (mean: 0.9 s per report) and the breast cohort in 2 min 36 s (mean: 4.6 s per report). DeepPhe-XN (v0.7.0) was executed locally in a Docker container on Red Hat Enterprise Linux 8.9 with 2 logical CPU cores and a 16 GB memory limit. The container used Eclipse Temurin 8 for Java with a configured maximum heap size of 3 GB (-Xmx3G). DeepPhe-XN processed the pancreatic cohort in 6 min 4 s (mean: 1.1 s per report) and the breast cohort in 2 min 0 s (mean: 3.5 s per report). Runtimes reflect end-to-end wall-clock time, including file I/O; mean seconds per report were computed as total cohort runtime divided by the number of reports.

The two-iteration optimization protocol in Brim Analytics exploration yielded substantial performance improvements. Mean accuracy on the development cohort improved from 85.9% (95% CI [83.2, 88.6]) at Iteration 0 to 92.1% (95% CI [90.0, 94.2]) at Iteration 1, representing a 6.2 percentage point optimization gain (P < 0.001, paired comparison). As shown in Table 2, individual variable gains ranged from 3.0 to 8.0 percentage points.

**Table 2.** Brim Analytics Variable Performance During Iterative Development (n = 150)

| Variable | Iteration 0 Accuracy | Iteration 1 Accuracy | Gain (p.p.) |
| --- | --- | --- | --- |
| T stage | 85.0 | 92.0 | +7.0 |
| N stage | 95.0 | 98.0 | +3.0 |
| M stage | 96.0 | 99.0 | +3.0 |
| OncoTree | 84.0 | 92.0 | +8.0 |
| Tumor site | 78.0 | 86.0 | +8.0 |
| Margin status | 81.0 | 89.0 | +8.0 |
| Histologic grade | 82.0 | 89.0 | +7.0 |

Note. Iteration 0 = guideline-derived specifications without data exposure; Iteration 1 = specifications refined through systematic error analysis; p.p. = percentage points.

Descriptive pathologic variables showed the largest optimization gains. Tumor site had the lowest Iteration 0 performance (78.0%), largely due to anatomic terminology variation—pancreatic subsites were described with diverse synonyms (“head” vs. “cephalic portion” vs. “proximal pancreas”). An 8.0 percentage point improvement after systematic synonym expansion indicates that explicit terminology mapping can address this documentation of heterogeneity. Margin status and histologic grade similarly improved by 8.0 and 7.0 percentage points, respectively, after adding explicit rules for ambiguous qualifiers.

### Pancreatic Adenocarcinoma Validation Performance

A total of 330 pancreatic adenocarcinoma reports were analyzed, spanning 2009–2025, and encompassing heterogeneous report formats (72.1% narrative, 27.9% synoptic). Brim Analytics achieved high accuracy across all seven variables, with accuracy ranging from 90.6% to 99.7% (Table 3). For T stage, Brim Analytics correctly classified 318 of 330 cases (96.4%), achieving an F1 score of.977 and κ of.885—indicating near-perfect agreement with gold-standard abstractions. This performance substantially exceeded the 83.6% accuracy observed for DeepPhe on the same variable, where high recall (97.5%) was offset by lower precision (82.7%), indicating systematic overclassification of tumor extent.

**Table 3.** Performance Metrics for Pancreatic Adenocarcinoma Validation Cohort (N = 330)

| Variable | Platform | Accuracy (95% CI) | Precision | Recall | F1 | $\kappa$ |
| --- | --- | --- | --- | --- | --- | --- |
| T stage | Brim | 96.4% [93.8–97.9] | 99.6% | 95.9% | .977 | .885 |
|  | DeepPhe | 83.6% [79.3–87.2] | 82.7% | 97.5% | .895 | .536 |
| N stage | Brim | 95.8% [93.0–97.5] | 96.7% | 98.7% | .977 | .744 |
|  | DeepPhe | 96.4% [93.8–97.9] | 97.7% | 96.9% | .973 | .917 |
| M stage | Brim | 99.7% [98.3–99.9] | 99.7% | 100% | .998 | —<sup>a</sup> |
|  | DeepPhe | 91.5% [88.0–94.1] | 51.1% | 82.8% | .632 | .587 |
| OncoTree | Brim | 90.6% [87.0–93.3] | 94.6% | 95.5% | .951 | —<sup>a</sup> |
| Tumor site | Brim | 99.4% [97.8–99.8] | 99.4% | 100% | .997 | .947 |
| Margin status | Brim | 98.2% [96.1–99.2] | 98.5% | 99.7% | .991 | —<sup>a</sup> |
| Histologic grade | Brim | 97.0% [94.5–98.3] | 100% | 96.0% | .980 | .920 |
| **Mean** | **Brim** | **96.7%** | **98.4%** | **98.0%** | **.981** | **.874** |
|  | **DeepPhe** | **90.5%** | **77.2%** | **92.4%** | **.833** | **.680<sup>c</sup>** |

*Note.* Mean accuracy is the unweighted average across variables shown (macro-average). DeepPhe extracted TNM variables only; DeepPhe means therefore summarize TNM only. CI, confidence interval; $\kappa$ , Cohen's kappa coefficient. <sup>a</sup> $\kappa$ suppressed owing to small sample size and class imbalance. <sup>b</sup>Mean $\kappa$ not calculated owing to insufficient valid estimates. <sup>c</sup>Interpret with caution given sample size limitations.

N stage was the only domain where DeepPhe matched or slightly exceeded Brim Analytics (96.4% vs. 95.8%, Δ = +0.6 percentage points), with both platforms achieving F1 scores >.97. This suggests lymph node staging terminology may be more consistently represented in existing oncology ontologies. For M stage, Brim Analytics achieved near-perfect accuracy (99.7%), whereas DeepPhe showed markedly lower precision (51.1%) despite reasonable recall (82.8%), yielding an F1 of.632 and indicating a high false-positive metastatic classification rate.

Among descriptive variables evaluated using Brim Analytics, tumor site achieved the highest accuracy (99.4%, F1 =.997), benefiting from standardized anatomic terminology in both narrative and synoptic formats. Margin status (98.2%) and histologic grade (97.0%) similarly demonstrated robust performance despite the greater narrative heterogeneity characteristic of these data elements. OncoTree classification achieved the lowest accuracy (90.6%), reflecting the complexity of mapping free-text histologic descriptions to hierarchical ontology codes—though this performance nonetheless exceeds thresholds typically required for automated pre-population of registry fields.

### Breast Cancer Validation Performance

Cross-disease transfer to breast cancer revealed marked differences in platform generalizability (Table 4). Brim Analytics remained stable, with mean accuracy of 93.7%—only 3.0 percentage points lower than pancreatic performance—despite differences in anatomic terminology, staging conventions, and histologic classification systems. T stage achieved perfect classification (100%), a 3.6 percentage point improvement over pancreatic validation, likely due to more standardized tumor size reporting in breast pathology. Margin status also reached 100% accuracy, and histologic grade remained strong (97.1%) despite the use of a different grading system (Nottingham vs. WHO pancreatic criteria).

**Table 4.** Performance Metrics for Breast Cancer Validation Cohort (N = 34)

| Variable | Platform | Accuracy (95% CI) | Precision | Recall | F1 | $\kappa$ |
| --- | --- | --- | --- | --- | --- | --- |
| T stage | Brim | 100% [89.8–100] | 100% | 100% | 1.00 | —<sup>a</sup> |
|  | DeepPhe | 70.6% [53.8–83.2] | 71.9% | 95.8% | .821 | .076 |
| N stage | Brim | 94.1% [80.9–98.4] | 100% | 94.1% | .970 | —<sup>a</sup> |
|  | DeepPhe | 94.1% [80.9–98.4] | 96.8% | 96.8% | .968 | .634 |
| M stage | Brim | 88.2% [73.4–95.3] | 90.9% | 96.8% | .937 | —<sup>a</sup> |
|  | DeepPhe | 85.3% [69.9–93.6] | 50.0% | 20.0% | .286 | .220 |
| OncoTree | Brim | 82.4% [66.5–91.7] | 84.8% | 96.6% | .903 | —<sup>a</sup> |
| Tumor site | Brim | 94.1% [80.9–98.4] | 97.0% | 97.0% | .970 | —<sup>a</sup> |
| Margin status | Brim | 100% [89.8–100] | 100% | 100% | 1.00 | —<sup>a</sup> |
| Histologic grade | Brim | 97.1% [85.1–99.5] | 97.1% | 100% | .985 | —<sup>a</sup> |
| **Mean** | **Brim** | **93.7%** | **95.7%** | **97.8%** | **.966** | —<sup>b</sup> |
|  | **DeepPhe** | **83.3%** | **72.9%** | **70.9%** | **.692** | **.310<sup>c</sup>** |

*Note.* Mean accuracy is the unweighted average across variables shown (macro-average). DeepPhe extracted TNM variables only; DeepPhe means therefore summarize TNM only. CI, confidence interval; $\kappa$ , Cohen's kappa coefficient. <sup>a</sup> $\kappa$ suppressed owing to small sample size and class imbalance. <sup>b</sup>Mean $\kappa$ not calculated owing to insufficient valid estimates. <sup>c</sup>Interpret with caution given sample size limitations.

M stage exhibited the largest cross-disease decline for Brim Analytics (88.2%, Δ = −11.5 p.p.), likely reflecting differences in metastatic status documentation conventions between breast and pancreatic pathology reports. OncoTree classification accuracy decreased from 90.6% to 82.4% (Δ = −8.2 p.p.)—a predictable outcome given that variable optimization during Iterations 0 and 1 was performed exclusively on pancreatic adenocarcinoma cases, thereby encoding pancreatic-specific histologic terminology mappings. This performance gap represents a tractable engineering problem rather than a fundamental limitation: the iterative refinement protocol yielded an 8.0 percentage point improvement for OncoTree in pancreatic cancer (Table 2), and equivalent disease-specific optimization would be expected to achieve comparable gains for breast cancer classification.

DeepPhe showed more pronounced cross-disease differences, with mean staging accuracy declining 7.2 percentage points (90.5% pancreatic vs. 83.3% breast). T stage performance differed substantially (70.6%, a 13.0 percentage point decline from pancreatic), with the near-zero κ (.076) indicating classification approaching chance levels. M stage extraction exhibited severe deficits (85.3% accuracy, F1 =.286), driven by only 20% recall—suggesting the system frequently failed to identify metastatic indicators in breast cancer documentation.

### Error Analysis

Error distribution analysis revealed systematic differences in platform failure modes (Table 5). Brim Analytics exhibited predominantly conservative error patterns, with T stage errors in pancreatic cancer showing strong false negative bias (11 FN vs. 1 FP). This conservative tendency—underclassifying rather than overclassifying tumor extent—is likely clinically preferable, especially in a human-in-the-loop setting, because false positive staging could inappropriately upstage patients. In contrast, DeepPhe demonstrated heavily skewed false positive errors for T stage (48 FP vs. 6 FN in pancreatic; 9 FP vs. 1 FN in breast), indicating systematic overclassification that persisted across disease contexts.

**Table 5.** Error Distributions by Variable, Platform, and Disease.

| Variable | Pancreas |  | Breast |  |
| --- | --- | --- | --- | --- |
|  | Brim, n (%) | DeepPhe, n (%) | Brim, n (%) | DeepPhe, n (%) |
| T stage | 12 (3.6) [1/11] | 54 (16.4) [48/6] | 0 (0.0) [0/0] | 10 (29.4) [9/1] |
| N stage | 14 (4.2) [10/4] | 12 (3.6) [5/7] | 2 (5.9) [0/2] | 2 (5.9) [1/1] |
| M stage | 1 (0.3) [1/0] | 28 (8.5) [23/5] | 4 (11.8) [3/1] | 5 (14.7) [1/4] |
| OncoTree | 31 (9.4) [17/14] | — | 6 (17.6) [5/1] | — |
| Tumor site | 2 (0.6) [2/0] | — | 2 (5.9) [1/1] | — |
| Margin status | 6 (1.8) [5/1] | — | 0 (0.0) [0/0] | — |
| Histologic grade | 10 (3.0) [0/10] | — | 1 (2.9) [1/0] | — |

Note. Brackets indicate [false positives/false negatives]; em dashes indicate variable not extracted by DeepPhe.

Stratification by report format revealed that Brim Analytics demonstrated format-agnostic performance (free-text error rate 4.6% vs. synoptic 1.1% in pancreatic cancer), with error distributions proportional to cohort composition. DeepPhe exhibited disproportionately elevated error rates on narrative reports (21.4% vs. 3.3% for synoptic in pancreatic cancer), indicating strong dependence on structured documentation that substantially limits utility for legacy or narrative-predominant report archives (Table 6).

**Table 6.**
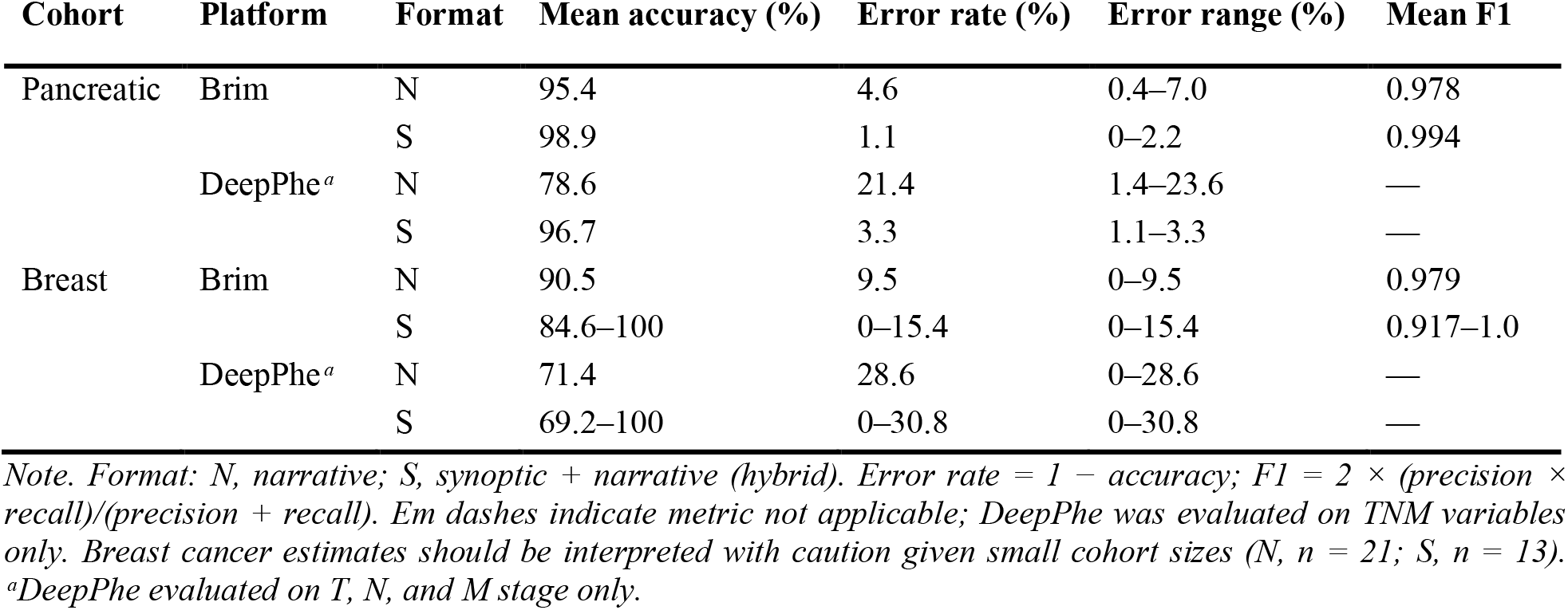
Pathology report extraction performance by platform, cancer type, and report format.

## Discussion

Our study demonstrates that contemporary AI-driven NLP platforms can extract critical pathology variables from unprocessed clinical reports with clinically meaningful accuracy. Brim Analytics achieved high accuracy across all variables (mean 96.7% for pancreatic cancer, 93.7% for breast cancer), with TNM staging accuracy exceeding 95% for pancreatic cancer. Minimal cross-disease degradation (3.0 percentage points) suggests that explicit, guideline-based extraction rules grounded in shared clinical frameworks (AJCC criteria, CAP guidelines) generalize well across anatomically distinct malignancies—an important requirement for operational deployment where disease-specific optimization would be prohibitive. DeepPhe showed comparable N stage performance (96.4% pancreatic, 94.1% breast) but substantial T stage deficits (83.6% pancreatic, 70.6% breast). The 12.8–29.4 percentage point accuracy gap and consistent false-positive bias highlight challenges in ontology-based mapping of heterogeneous anatomic descriptors. The larger cross-disease decline (7.2 percentage points) may also reflect limited ontology coverage for breast-specific terminology. These findings extend prior work^9,10^ by systematically evaluating cross-disease generalizability under deployment-realistic conditions.

Three limitations warrant consideration. First, this single-institution evaluation may not generalize to documentation practices at other healthcare systems; future work should include multi-institutional validation. Second, the breast cancer cohort (n = 34) limited statistical power to detect moderate performance differences, and additional validation rounds are needed to assess operational readiness. Third, neither platform was customized or integrated into a hybrid pipeline, nor did they evaluate disease-specific optimization. Future work should also include formal cost-effectiveness analyses comparing automated versus manual abstraction.

The architectural differences between the two platforms carry significant operational implications that extend beyond raw accuracy and contextualize where our findings sit relative to prior work. These results are broadly consistent with previous NLP-based pathology extraction studies, though direct comparisons are limited by differences in cancer type, variable scope, and ground truth methodology. For example, Hochheiser^12^ evaluated the DeepPhe-CR system across five cancer types from two SEER registries, reporting pathological TNM F1 scores of 0.94–0.99, with broader histomorphological attributes ranging from 0.81 to 0.96, consistent with the synoptic-format performance observed here. These were notably higher TNM F1 scores than the free-text performance observed in our pancreatic cohort, further reinforcing the format-related performance differential reported here. Moreover, it should be noted that pancreatic staging carries unique complexity, including vascular involvement, margin status, and perineural invasion, that is not captured in this comparator cohort; these figures should therefore be interpreted as a nearest available approximation rather than a direct equivalent.

Across both cancer types, our study extends the existing literature by systematically stratifying extraction performance by report format, a dimension that prior benchmarking studies have not addressed. Free-text error rates exceeded synoptic error rates by 4.2-fold for Brim Analytics and 6.5-fold for DeepPhe in pancreatic cancer, demonstrating that report format is an underappreciated but consequential source of performance variability. This distinction has direct implications for institutional deployment. Brim Analytics’ explicit, human-readable extraction rules enable transparent auditing and rapid adaptation to evolving guidelines without model retraining, but require domain expert involvement to develop and refine variable specifications. DeepPhe’s ontology-driven approach provides out-of-the-box functionality with minimal configuration, but may necessitate substantial ontology engineering when extending to new disease domains or extraction targets not covered by existing cancer phenotype models. Both platforms support human-in-the-loop workflows, though their error characteristics differ: Brim Analytics exhibited predominantly false-negative errors (underclassification), while DeepPhe showed false-positive bias for certain variables, particularly T stage. Processing speeds for both platforms (0.9–4.6 s/report) are operationally equivalent, indicating that platform selection should be guided by institutional factors including available informatics expertise, tolerance for upfront configuration effort, requirements for extraction rule transparency, and sensitivity to specific error types in downstream workflows.

Beyond technical feasibility, these findings point toward a broader role for automated NLP extraction within integrated cancer data ecosystems. Cancer centers increasingly operate under data governance frameworks that require timely, standardized, and auditable abstraction of clinical variables across large patient volumes. In this context, platforms could function not as replacements for certified tumor registrars, but as a first-pass abstraction layer that pre-populates registry fields, prioritizes cases requiring human review, and flags discrepancies between structured and narrative report content. This human-in-the-loop model, in which automation handles high-confidence extractions and escalates ambiguous cases, is consistent with the computer-assisted abstraction paradigm demonstrated by DeepPhe and would be practically achievable with current platform performance levels for synoptic reports. Federated research networks, such as AACR Project GENIE, which aggregate multi-institutional genomic and clinical data at scale, represent a particularly compelling deployment context. For example, automated extraction of TNM staging and histopathological variables across participating sites could accelerate case-level data harmonization, reduce inter-institutional abstraction variability, and expand the volume of analytically ready data available for real-world evidence generation — provided that platform performance is validated across the documentation practices of contributing institutions.

The immediate next steps for this line of work are threefold. First, multi-institutional validation across health systems with diverse documentation cultures and EHR platforms is essential to establish generalizability beyond the single-center cohort evaluated here. The recently established Cancer AI Alliance (CAIA) of top cancer centers (SKCCC is one of them) offers an opportunity for pipeline sharing across participating institutions and may support such validations^18^. Second, prospective integration studies embedding automated extraction within live registry workflows would enable direct measurement of efficiency gains, error-detection rates, and registrar acceptance—outcomes that benchmarking studies alone cannot address. Third, expansion to additional cancer types with complex staging architectures, including lung, colorectal, and prostate cancer, would test whether the cross-disease stability observed here for Brim Analytics persists at scale. Together, these steps would build the evidence base needed for policy-level adoption of automated abstraction in national cancer surveillance infrastructure and clarify when full or partial automation becomes operationally warranted.

## Data Availability

All data produced in the present study are available upon reasonable request to the authors.

## Acknowledgements

This work was supported in part by the National Cancer Institute (grant U01CA274631). We thank Daniel Fabbri (Vanderbilt University) for assistance with BRIM Analytics; Sean Finan (Boston Children’s Hospital), John Levander (University of Pittsburgh), and Guergana Savova (Boston Children’s Hospital and Harvard Medical School) for assistance with DeepPhe.

## References

1. Zachary I, Boren SA, Simoes E, Jackson-Thompson J, Davis JW, Hicks L. Information management in cancer registries: Evaluating the needs for cancer data collection and cancer research. Online J Public Health Inform. 2015;7(3):e213. doi:10.5210/ojphi.v7i3.6170

2. Siegel RL, Miller KD, Wagle NS, Jemal A. Cancer statistics, 2023. CA Cancer J Clin. 2023;73(1):17–48. doi:10.3322/caac.21763

3. Hofferkamp J. Standards for Cancer Registries Volume III: Standards for Completeness, Quality, Analysis, and Management of Data. Springfield, IL: North American Association of Central Cancer Registries; 2011.

4. Yim WW, Yetisgen M, Harris WP, Kwan SW. Natural language processing in oncology: A review. JAMA Oncol. 2016;2(6):797–804. doi:10.1001/jamaoncol.2016.0213

5. Kreimeyer K, Foster M, Pandey A, Arya N, Halford G, Jones SF, Forshee R, Walderhaug M, Botsis T. Natural language processing systems for capturing and standardizing unstructured clinical information: A systematic review. J Biomed Inform. 2017;73:14–29. doi:10.1016/j.jbi.2017.07.012

6. Kreimeyer K, Barasa D, Sherief M, Shi X, Borja M, Yegnasubramanian S, Anagnostou V, Murray JC, Botsis T. An implemented real-world-data pipeline for standardization of electronic health records in precision oncology. AMIA Jt Summits Transl Sci Proc. 2025;2025:242–249.

7. Singhal K, Azizi S, Tu T, Mahdavi SS, Wei J, Chung HW, Scales N, Tanwani A, Cole-Lewis H, Pfohl S, Payne P, Seneviratne M, Gamble P, Kelly C, Babiker A, Schärli N, Chowdhery A, Mansfield P, Demner-Fushman D, Agüera Y Arcas B, et al. Large language models encode clinical knowledge. Nature. 2023;620(7972):172–180. doi:10.1038/s41586-023-06291-2

8. Savova GK, Masanz JJ, Ogren PV, Zheng J, Sohn S, Kipper-Schuler KC, Chute CG. Mayo clinical text analysis and knowledge extraction system (cTAKES): Architecture, component evaluation and applications. J Am Med Inform Assoc. 2010;17(5):507–513. doi:10.1136/jamia.2009.001560

9. Alawad M, Gao S, Qiu JX, Yoon HJ, Blair Christian J, Penberthy L, Mumphrey B, Wu XC, Coyle L, Tourassi G. Automatic extraction of cancer registry reportable information from free-text pathology reports using multitask convolutional neural networks. J Am Med Inform Assoc. 2020;27(1):89–98. doi:10.1093/jamia/ocz153

10. Gao S, Young MT, Qiu JX, Yoon HJ, Christian JB, Fearn PA, Tourassi GD, Ramanthan A. Hierarchical attention networks for information extraction from cancer pathology reports. J Am Med Inform Assoc. 2018;25(3):321–330. doi:10.1093/jamia/ocx131

11. Brim Analytics. AI-guided medical chart abstraction. Accessed July 10, 2025. https://www.brimanalytics.com/

12. Hochheiser H, Finan S, Yuan Z, Durbin EB, Jeong JC, Hands I, Rust D, Kavuluru R, Wu XC, Warner JL, Savova G. DeepPhe-CR: Natural language processing software services for cancer registrar case abstraction. medRxiv. 2023. doi:10.1101/2023.05.05.23289524

13. Savova GK, Tseytlin E, Finan S, Castine M, Miller T, Medvedeva O, Harris D, Hochheiser H, Lin C, Chavan G, Jacobson RS. DeepPhe: A natural language processing system for extracting cancer phenotypes from clinical records. Cancer Res. 2017;77(21):e115–e118. doi:10.1158/0008-5472.CAN-17-0615

14. Botsis T, Murray JC, Ghanem P, Balan A, Kernagis A, Hardart K, He T, Spiker J, Kreimeyer K, Tao J, Baras AS, Yegnasubramanian S, Canzoniero J, Anagnostou V; Johns Hopkins Molecular Tumor Board Investigators. Precision oncology core data model to support clinical genomics decision making. JCO Clin Cancer Inform. 2023;7:e2200108. doi:10.1200/CCI.22.00108

15. Kundra R, Zhang H, Sheridan R, Sirintrapun SJ, Wang A, Ochoa A, Wilson M, Gross B, Sun Y, Madupuri R, Satravada BA, Reales D, Vakiani E, Al-Ahmadie HA, Dogan A, Arcila M, Zehir A, Maron S, Berger MF, Viaplana C, et al. OncoTree: A cancer classification system for precision oncology. JCO Clin Cancer Inform. 2021;5:221–230. doi:10.1200/CCI.20.00108

16. McHugh ML. Interrater reliability: The kappa statistic. Biochem Med. 2012;22(3):276–282. doi:10.11613/BM.2012.031

17. Newcombe RG. Two-sided confidence intervals for the single proportion: Comparison of seven methods. Stat Med. 1998;17(8):857–872. doi:10.1002/(SICI)1097-0258(19980430)17:8<857::AID-SIM777>3.0.CO;2-E

18. Cancer AI Alliance. Cancer AI Alliance (CAIA): Accelerating AI for cancer research. Accessed July 10, 2025. https://www.canceralliance.ai/

